# Establishing Biomarkers and Clinical Endpoints in Myotonic Dystrophy Type 1 (END-DM1): protocol of an international natural history study

**DOI:** 10.1101/2025.08.13.25333647

**Authors:** Karlien Mul, Kate Eichinger, Man Hung, Valeria A. Sansone, Cynthia Gagnon, Sub Subramony, Richard H. Roxburgh, Johanna Hamel, Jeffrey M. Statland, Bakri Elsheikh, Chris Turner, Jacinda Sampson, Thomas Ragole, Emma Matthews, Benedikt Schoser, Andrea Swenson, Chamindra Laverty, Perry Shieh, Ericka P. Greene, Masanori Takahashi, Matthew Wicklund, Jeanne Dekdebrun, Jennifer Raymond, Erin DeSpain, Charles A. Thornton, Nicholas E. Johnson, Myotonic Dystrophy Clinical Research Network

## Abstract

**Background:** Myotonic dystrophy type 1 (DM1) is an autosomal dominant inherited multi-system disorder that affects skeletal muscles but also many other organ systems. Molecular targets have been identified and targeted therapies are being developed and tested in first-in-human clinical trials. However, insufficient knowledge of the phenotypic heterogeneity and natural course of the disease, together with a lack of reliable biomarkers, complicate the design of clinical trials.

**Methods:** The main objectives of this study are to 1) characterize the phenotypic heterogeneity and disease progression of DM1 in a large cohort; 2) identify baseline characteristics that predict subsequent progression; 3) validate RNA biomarkers of disease severity. This is a prospective, multi-site observational study with a follow-up period of 24 months including approximately 700 adult DM1 patients. Visits will occur at baseline, and months 12 and 24. All patients will undergo strength testing, myotonia assessment, a battery of functional outcome assessments, spirometry, and complete various questionnaires and cognitive tests. Blood and urine samples will be taken at each visit for biomarker studies. A subset of 60 patients will undergo a muscle biopsy at baseline and at an additional 3-month visit.

The sensitivity to disease progression and minimally clinically important differences will be determined for the various clinical outcome measures. Associations between baseline patient characteristics and the rate of disease progression will be evaluated.

**Discussion:** The results of this large international study on DM1 will contribute to optimizing clinical trial design. Both data and biological samples will be collected for future research as well.

**Trial registration:** clinicaltrials.gov NCT03981575

## Background

Myotonic dystrophy type 1 (DM1) is an autosomal dominant inherited disease that affects many organs and reduces life expectancy. In addition to prominent muscular symptoms, DM1 also has important smooth muscle, central nervous system, endocrine, ocular and cardiac manifestations. Its prevalence is estimated at 9 per 100,000, but varies widely in different regions throughout the world.(1) Currently, there is no cure or treatment available to slow disease progression. Patient care is aimed at symptomatic management and preventing and treating disease complications.

DM1 is caused by a trinucleotide expansion of the CTG repeat in the *dystrophia myotonica protein kinase* (*DMPK)* gene on chromosome 19.(2, 3) A repeat expansion of at least 50 repeats leads to sequestration of RNA-binding proteins, resulting in dysregulated alternative splicing for many genes. Although pathways for RNA gain-of-function are not fully known, a consensus has been reached that RNA toxicity is the core mechanism for disease.(4)

With the discovery of this novel disease process, molecular targets have been identified and targeted therapies are being developed.(4) Several candidate therapeutics have already advanced to first-in-human clinical trials. However, there are still hurdles to overcome to optimize the design of clinical trials and determine what would be clinically meaningful changes for patients on new investigational products.

DM1 is highly variable in terms of the age at symptom onset, the pattern of organ involvement, and the extent of disease severity and progression. Given this heterogeneity, large studies and long duration may be required to show treatment effects in trials, and detect dose-response relationships. Potential strategies to improve trial efficiency include selecting the patients most likely to progress during a defined interval, or stratifying the allocation based on estimated trajectory of progression. However, apart from the well-known relationship between longer repeat length and younger symptom onset and greater severity, currently there is no rational basis to make these selections or perform stratification.(5) Accordingly, one of the aims of this study is to expand knowledge regarding predictors of disease progression.

Equally important is the development of robust biomarkers to guide drug development and provide confirmatory evidence of drug activity in clinical trials. Earlier work has shown that splicing dysregulation is a promising biomarker that is highly responsive to treatment in animal models. However, while bulk RNA sequencing is an excellent tool for discovering the splicing defects in DM1, its application for repeated sampling in large cohorts, using small biopsy samples, is problematic. This has triggered efforts to develop high-precision, higher-throughput methods for analyzing key splice events. Accordingly, this study will also assess biopsy-rebiopsy variability and longitudinal changes for these methods.

Historically there have been few multicenter controlled trials in DM1, but this is changing rapidly as many lines of drug development converge on clinical testing. This places a premium on conducting trials in the most efficient manner, so that many candidate therapies can be evaluated in parallel. To this end, the END-DM1 study was also initiated to chart a path towards definitive and highly efficient trials. Here we describe the objectives and methods of this international, prospective, observational study.

## Objectives

The overarching goal of this study is to overcome obstacles in the design and conduct of clinical trials for newly developed therapies in DM1.

The primary objectives of the study are:

1. To characterize the phenotypic heterogeneity and disease progression of DM1 in a large cohort;
2. To identify baseline characteristics that predict subsequent progression;
3. To further validate RNA biomarkers of disease severity; and
4. To develop a group of centers and investigators who are experienced with outcome measures and procedures for the DM1 population and committed to collaborative research to establish new treatments.

## Methods/design

### Design

The END-DM1 study is a prospective, multi-site observational study with a follow-up period of 24 months. The study is registered at clinicaltrials.gov (NCT03981575). Recruitment started in October 2018, was impacted by the Covid pandemic, and is currently ongoing.

Participant recruitment is expected to be completed by Q3 2025, with final data collection concluding in 2027. Baseline analyses will be conducted and published during the study, while complete longitudinal results are anticipated from 2027 onwards.

### Participant timeline

The SPIRIT schedule (fig 1) summarizes the time points for enrolment and all study procedures, including an additional month 3 visit for 60 participants in the muscle biopsy subgroup. Visits may be carried out over 2 days as long as the second day occurs within 7 days. End of Study visits will be allowed out of window to allow for more complete data collection. Missed visits can be made up out of window.

**Figure 1.**
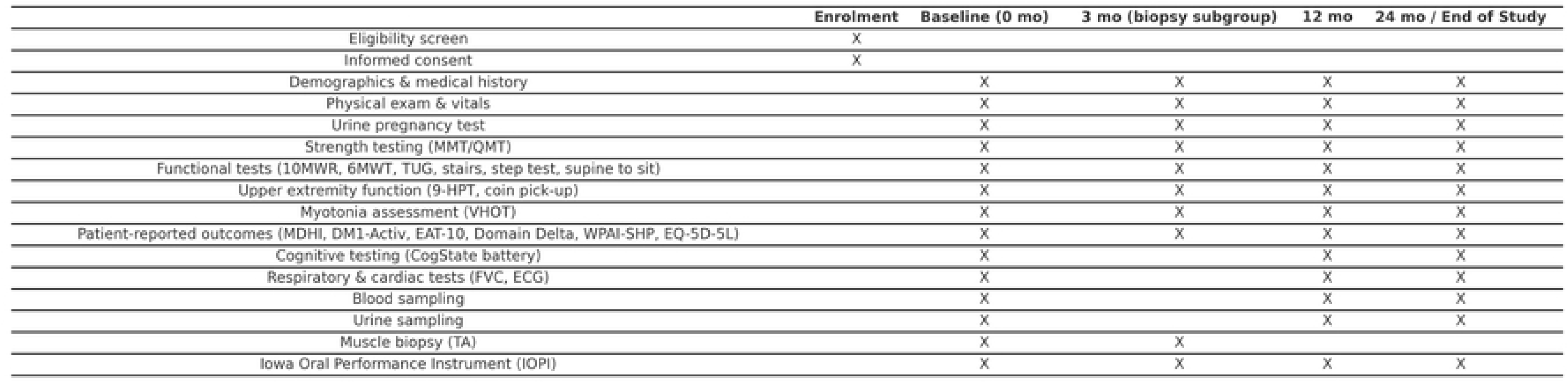
SPIRIT schedule of enrolment and assessments for the END-DM1 study. No interventions are planned in this observational natural history study; the schedule outlines enrolment and assessment time points only. “X” indicates that the procedure is performed at the given time point. The baseline visit is at month 0, with follow-up visits at months 12 and 24 for all participants, and an additional visit at month 3 for 60 participants in the muscle biopsy subgroup.

### Study setting

Approximately 700 patients will be recruited from 22 centers around the world (fig 2). The participating centers are part of the Myotonic Dystrophy Clinical Research Network (DMCRN), an international consortium of investigators who are focused on DM1.

**Figure 2.**
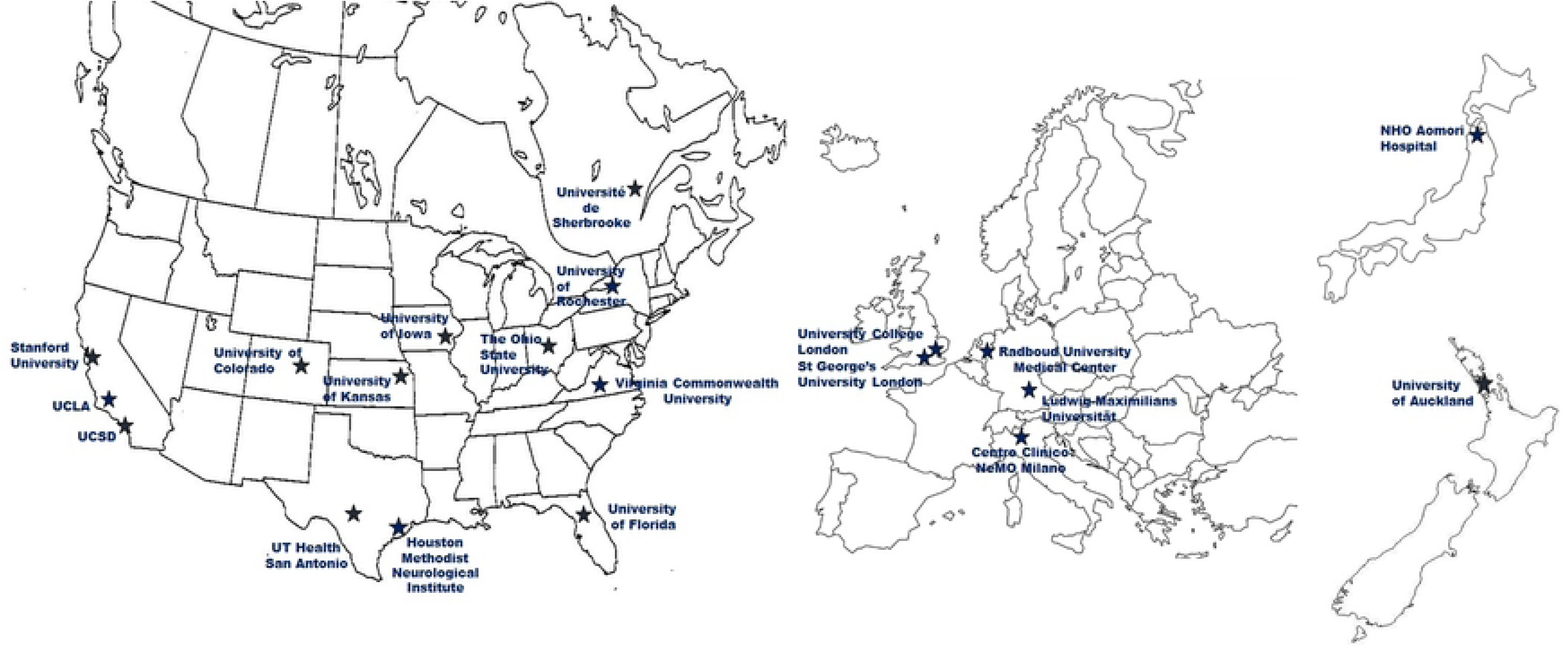
The DMCRN is a multinational consortium conducting the END-DM1 study.

### Ethics approval and consent to participate

For each participating center the protocol and informed consent forms were approved by an ethics committee and relevant regulatory authorities. The END-DM1 study was reviewed by WIRB-Copernicus Group Institutional Review Board at the study level and at the site level for the majority of sites in the United States (reference number 20204399, initial approval on Dec 13^th^ 2017). Each site performed a local context review to adjust the informed consent to address any local considerations. Sites outside of the US had protocol reviewed by their separate institution human subject committees. For the study site in Milan, Italy, the ethics review committee Comitato Etico Milano Area 3 reviewed and approved the study (reference number ID 4797). For the study site in Nijmegen, Netherlands, the ethics review committee CMO Arnhem-Nijmegen Region reviewed and approved the study (reference number NL.71565.091.20). For the study site in Auckland, New Zealand, the Central Health and Disability Ethics Committee reviewed and approved the study (reference number 2024 AM 11251). For the study site in Sherbrooke, Canada, the ethics review committee Le Comité d’éthique de la recherche du Centre intégré universitaire de santé et de services sociaux du Saguenay-Lac-Saint-Jean reviewed and approved the study (reference number 2022-055). For the study site in London, United Kingdom, the ethics review committee London – South East Research Ethics Committee reviewed and approved the study (reference number 282941). For the study site in Munich, Germany, the ethics review committee Ethik-Kommission bei der Medizinischen Fakultät der Ludwig-Maximilians Universität München reviewed and approved the study (reference number 20-0822). For the study site in Osaka, Japan, the ethics review committee Osaka University Research Ethics Committee reviewed and approved the study (reference number 23469(T1)-2).

The study is being conducted under the Declaration of Helsinki and Good Clinical Practice guidelines, which include the protection of data and patient privacy. All participants are required to provide written informed consent before participating.

### Population

Patients that fulfill the following criteria are eligible to participate in the study: Inclusion criteria:

- A clinical diagnosis of DM1 based on research criteria,(6) or genetic confirmation of DM1 diagnosis
- Adult patients ages 18 to 70 years old (inclusive)
- Competent to provide informed consent

Exclusion criteria:

- Symptomatic renal or liver disease, uncontrolled diabetes or thyroid disorder, or active malignancy other than skin cancer
- Current alcohol or substance abuse
- Concurrent enrollment in clinical trial for DM1, or participation in trial within 6 months of entry
- Concurrent pregnancy or planned pregnancy during the course of the study
- Concurrent medical condition that would, in the opinion of the investigator or clinical evaluator compromise performance on study measures
- Use of mexiletine or other anti-myotonia agents within 72 hours of baseline visit
- Note: non-ambulatory participants are not excluded but are limited to <15% of enrollment

At least half of the 60 patients undergoing the tibialis anterior (TA) muscle biopsy will need to have moderate or severe weakness of ankle dorsiflexion, defined as MRC score ≤ 4+, to mimic expected populations in clinical trials, and drive robust comparison of muscle weakness with splicing defects. For this subgroup there are additional exclusion criteria, namely:

- CTG repeat expansion size less than 80 repeats, unless there are clear cut signs of limb weakness and muscle wasting
- Conditions or medication that increase the risk of bleeding, including anticoagulant use, history of bleeding or a known platelet count < 50 × 10^9^/L
- Advanced wasting of TA muscle that, in the opinion of the investigator, makes it unlikely that needle biopsy can obtain an adequate muscle sample
- Previous muscle biopsy of either TA, in order to provide muscle tissue samples of non-biopsied muscles

### Procedures and assessments

Prior to enrollment, written informed consent will be obtained from every participant. Study visits will be performed at baseline and months 12 and 24. A subset of 60 participants has an extra visit 3 months after baseline and undergoes a muscle biopsy at baseline and the 3 month visit in addition to the standard set of assessments. An overview of assessments at each visit is provided in figure 1. Study visits can take place over 2 days, provided that day 2 is within 7 days of day 1. Assessments will be performed in a fixed order by trained physical therapists serving as clinical evaluators.

At baseline, demographic and social status information such as sex, race, ethnicity, education, and employment status will be collected. A medical history and physical examination will be performed and vital signs will be recorded. The age at onset of various symptoms will be documented, as well as common comorbidities.

Forced vital capacity (in sitting and supine position) and electrocardiograms will be performed to assess respiratory and cardiac involvement. Measures of muscle strength and timed function will be performed to assess impact on skeletal muscle. Patient reported outcome measures will be used to assess overall impact across many organ systems and extend of neurologic impairment, as described below.

#### Strength testing

Strength measurements will include manual muscle testing (MMT) of neck flexors and extensors and 13 bilateral muscle groups: shoulder abductors, elbow flexors and extensors, wrist flexors and extensors, thumb flexors, hip flexors, extensors, and abductors, knee flexors and extensors, ankle dorsiflexors, and plantar flexors. The modified Medical Research Council (MRC) scale will be used to quantify strength in these groups.(7)

Quantitative strength testing (QMT) will be performed using a fixed myometry system with a force transducer attached by an inelastic strap to a metal frame. QMT testing will include five muscle groups bilaterally: elbow flexors and extensors, knee flexors and extensors and ankle dorsiflexors.(8) Handgrip strength will measured using a digital dynamometer. Both MMT and QMT will be performed with standardized positions and direction of movement per muscle.

For orofacial strength the Iowa Oral Performance Instrument (IOPI) will be used to measure tongue elevation and bilateral cheek compression (‘buccal’) strength in kPa.(9) A bulb will be placed between the tongue and the palate or the cheek and the jaw respectively, and the participant will be asked to squeeze the bulb as hard as possible.

#### Functional performances: gait and mobility

Various functional performance tests will assess gait and mobility. For the 10-meter walk/run (10-MWR) test patients will be asked to go 10 meters as quickly as possible, whether by walking or running, and the time will be recorded. The 6-minute walk test (6-MWT) will record the distance covered over a time of 6 minutes on a 20-meter course.(10) The timed-up-and-go (TUG) test will measure the time that it takes a patient to rise from a chair, walk 3 meters, turn around, return to the chair, and sit.(11) The timed supine to sit will assess the time it takes a patient to move from supine to sitting on the edge of a plinth. For the timed 4-Stair-Climb and 4-Stair-Descend test patients will be asked to climb up and down 4 stairs as quickly as possible, using steps with a six-inch rise and ten-inch run. Quality grades will be used to record reliance on railings and manner of ascent (i.e., step-to-step or step-over-step pattern). The step test will have the patient maintain stationary balance on one leg while the other footsteps rapidly on and off a 7.5 cm step. The score is the number of repetitions completed in 15 seconds.

#### Myotonia assessment

The degree of myotonia will be quantified by the video hand opening time (VHOT).(12) To eliminate “warm-up” of myotonia the patient will have to rest the dominant hand for five minutes. The patient will then squeeze the hand for three seconds and when prompted to do so, will open the hand as quickly as possible. A video recording of the procedure will be obtained showing the forearm and hand but not the face. Recordings will be assessed afterwards by a blinded reviewer to determine the hand opening time.

#### Functional performances: upper extremity function

To assess upper extremity performance timed functional tests are used. The patient will have to pick up 6 coins and hold them in the hand as quickly as possible (coin picking). For the 9 hole peg test (9-HPT) the patient will place 9 pegs in the 9 holes on the board and then remove them as quickly as possible.(13)

#### Patient reported outcome measures

Various questionnaires will be administered in the patient’s native or fluent language. The Myotonic Dystrophy Health Index (MD-HI) will serve to measure the DM1 disease burden across 17 different domains.(14)

The DM1-activity and participation scale (DM1-Activ) is a Rasch-built interval scale that will follow-up on daily and social functioning of DM1 patients.(15)

The EAT-10 is a 10-item questionnaire designed to assess difficulties with swallowing or dysphagia.(16)

The Domain Delta questionnaire will evaluate patient-perceived changes by asking the patient on a 5-points scale whether overall symptoms or specific domains have become worse, remained stable, or improved.(17)

The Work Productivity and Activity Impairment Questionnaire: Specific Health Problem V2.0 (WPAI:SHP) is a brief questionnaire that will evaluate the impact of the disease on a patient’s ability to work.(18)

The EQ-5D will provide a generic measure of health by assessing five dimensions, each describing a different aspect of health, namely mobility, self-care, usual activities, pain / discomfort and anxiety / depression.(19)

The walking scale-12 questionnaire will evaluate walking ability.

#### Cognitive function

To evaluate cognitive function a series of seven tests from the CogState, a computer-assisted method of cognitive assessment, will be employed.(20) The Cogstate tests are designed specifically for repeated assessment. The selection of tests intends to capture executive dysfunction which has previously been described in some DM1 patients.(21) The complete set of tests will measure various cognitive domains including verbal learning (International Shopping List task), executive function (Groton Maze Learning test and Set-shifting test), psychomotor function (Detection test), working memory (One Back test), attention (Identification test) and emotional recognition (Social-Emotional Cognition test).

### Biological samples

At each visit, blood samples will be collected for genetic testing and for biomarker discovery purposes. Samples may be used to extract plasma, DNA, RNA, metabolites or proteins. Urine samples will be collected at each visit for biochemical analysis.

A subgroup of 60 patients will undergo muscle biopsies from the tibialis anterior muscle at the baseline and three-month visits. The initial biopsy will be from the left leg, the second biopsy from the right leg. The tissue will be obtained through a needle biopsy as described previously.(22) A total of 2-4 aspirations from the same site will be taken. The samples will be snap frozen in liquid nitrogen in a single vial and used for analyses of RNA, and potentially DNA, proteins, or histological analysis.

Participants in the study will provide their consent for muscle tissue, blood, and urine to be placed in a BioBank. Materials in the BioBank that remain after completion of this study may be used for future research by investigators within or outside this study group.

### Multi-site infrastructure

Virginia Commonwealth University will serve as the overall coordinating study, responsible for participant recruitment, project and data management, and statistical core. Study oversight will be the responsibility of the Executive committee, which is composed of the study co-investigators, the lead physical therapist (PT) clinical evaluator, and study biostatistician. An in-person formal site initiation meeting took place prior to the start of the study, where focus was directed towards training investigators, study coordinators, clinical evaluators (CE) and other relevant site staff. To help ensure proper and consistent test administration across evaluators/sites, all evaluators undergo training during the investigator meeting on the proper administration of clinical outcome assessments.

A yearly meeting will be organized throughout the conduct of the study to review the protocol, recruitment and challenges with achieving study goals, and refresher training on clinical assessments. Additionally, regular conference calls will be scheduled with participating sites.

### Availability of data and materials

Data and biological samples collected in this study will be made available to both academic and commercial parties upon reasonable request.

### Statistical analyses and power/sample size calculation

The change in outcome measures from baseline to 12 and 24 months will be tested for statistical significance by a two-tailed, paired t-test (standardized response mean). All statistical significance will be set at alpha of 0.05. Additionally, responsiveness will be assessed by determining the effect size.

The responsiveness-to-change index will be assessed by dividing the absolute change scores in the DM1 group with stratification at the subgroup level (e.g., age, gender). Go-no go decision point for each outcome measure will be set for a responsive index of at least 0.5. This decision will be reviewed by the study investigators and advisory panel periodically during the project period and adjusted as necessary.

Further statistical analyses will be mainly focused on demonstrating clinically relevant changes in clinical outcomes. For determining the ‘minimally clinically importance difference (MCID)’ both distributional and anchor-based methods will be used. The distributional method chosen is that of the standard error of measurement (SEM). The SEM is the variability in the results from a specific clinical outcome measures that is attributed to instrument unreliability, in which a change smaller than the calculated SEM is likely to be due to measurement error rather than a true change.(23) For the anchor-based method, clinical outcomes will be compared among the categories of the ‘Domain Delta’ questionnaire (e.g.: unchanged, a little better, a lot better, etc.) using the Jonckheere-Terpsta test.

During the conduct of the study interim analyses will be performed to direct efforts and identify domains that need to be further supported or, on the other hand, avoided. The sample size for this study is designed to capture a clinically meaningful change in the clinical outcome assessments. In order to estimate a sample size based on the continuous variable of change in time in the 10-meter walk/run, the following assumptions were made to capture precision: a mean change of 0.277 with a standard deviation of 1.00. With an alpha of 0.05 and a two-sided two samples t-test achieving 93.5% power, a total sample size of 700 subjects will be needed to compare two subgroups of 315 individuals each in the mild DM1 and the more severely affected DM1 groups. Separating these groups will be key to identifying those individuals most suited for clinical trials. This sample size accounts for a 10% drop out rate, as was identified in prior studies. This sample size would also allow for computation of sensitivity and specificity of the primary endpoints to discriminate these two subgroups.

## Discussion

The results from this large international natural history study on DM1 will provide useful information to guide the design and conduct of future clinical trials.

The natural history data will help us gauge which outcome measures, both performance-based and patient reported, are capable of detecting changes over a two-year time frame. Importantly, it will also provide an estimate of minimum changes that are considered clinically significant to patients.

We also expect to improve the understanding of the clinical heterogeneity in DM1 and identify a range on the primary endpoint that may stratify the population into a group that is most likely to exhibit disease progression during the course of a clinical trial.

There are limitations to this study. DM1 can affect many different organs. Not all possible symptoms and disease complications are having a detailed assessment in the study. We prioritized the assessments to focus on aspects that matter most to patients, are tractable for study visits in clinical trials, and likely to detect disease progression over one and two year intervals.

As patients have to be willing, able, and interested to travel to research sites for a study that does not involve a therapeutic intervention, the study cohort may not be representative of the entire DM1 population. There may be underrepresentation of more severely affected patients, for example patients with more severe muscle weakness that are non-ambulatory. Similarly, patients with more severe cognitive deficits, or behavioral changes such as apathy, are less likely to participate in the study. Another drawback is that this study will not enroll patients under the age of 18. However, there are a separate natural history studies ongoing to assess children with DM1.

Adult patients with disease onset at pediatric age will be included the study. This group of patients generally has a more rapid disease progression and different severity of organ involvement. Data analysis will have to verify the impact of this population depending on how represented they will be in the end. The same will be true for patients with late-onset disease that are more often at the mild end of the disease spectrum.

A strength of the study is the worldwide collaboration of many centers, laying the groundwork for future clinical trials and drug approvals in broad geographic areas. A short-coming, however, is that less-developed countries are not represented.

Another strength is the training amongst sites with lead physical therapists sharing knowledge and experience with local therapists and providing regular feedback, ensuring data quality and consistency across sites.

Altogether, this study will aid in optimizing and ultimately accelerating the drug development process for DM1.

## Data Availability

No datasets were generated or analysed during the current study. All relevant data from this study will be made available upon study completion.

## List of abbreviations

DM1: myotonic dystrophy type 1
DMPK: dystrophia myotonica protein kinase
MDCRN: Myotonic Dystrophy Clinical Research Network
MRC: medical research council
TA: tibialis anterior
MMT: manual muscle testing
QMT: quantitative muscle testing
IOPI: Iowa Oral Pressure Instrument
10-MWR: 10 meter walk run test
6-MWT: 6 minute walking test
TUG: timed up and go test
VHOT: video hand opening time
9-HPT: 9 hole peg test
MD-HI: Myotonic Dystrophy Health Index

## Authors’ contributions

NEJ and CAT designed the study, obtained funding, are supervising the execution of the study and data interpretation, and revised and approved the manuscript. KM supervises the execution of the study and wrote the manuscript. VS, CG, SS, RR, JH, JS, BE, CT, JS, TR, EM, BS, AS, CL, PS, EG, MT, MW supervise the execution of the study, helped design the study and revised and approved the manuscript. KE, MH, JD, JR, ED helped design the study and revised and approved the manuscript.

## Acknowledgements

Collaborating author names for ‘Myotonic Dystrophy Clinical Research Network’:

- Michela Nani, Centro Clinico NeMO, Milan, Italy
- Enrico Cossu, Centro Clinico NeMO, Milan, Italy
- Valentina Franchino, Centro Clinico NeMO, Milan, Italy
- Carola Ferrari Aggradi, Centro Clinico NeMO, Milan, Italy
- Alice Zanolini, Centro Clinico NeMO, Milan, Italy
- Giovanni Colacicco, Centro Clinico NeMO, Milan, Italy
- Carino Jennings, Virginia Commonwealth University, Richmond VA, USA
- Aileen Jones, Virginia Commonwealth University, Richmond VA, USA
- Justine Dolbec, Sherbrooke University, Sherbrooke, Canada
- Amelie Lemire, Sherbrooke University, Sherbrooke, Canada
- Alex Rodriguez, University of Florida College of Medicine, Gainesville, FL, USA
- Donovan Lott, University of Florida College of Medicine, Gainesville, FL, USA
- Sarah Nagar, The University of Auckland, Auckland, New Zealand
- Sarah Mollet, The University of Auckland, Auckland, New Zealand
- James Hilbert, University of Rochester Medical Center, Rochester NY, USA
- Nicole Koopman, University of Rochester Medical Center, Rochester NY, USA
- Kaylene Whited, University of Kansas Medical Center, Kansas City KS, USA
- Sandhya Sasidharan, University of Kansas Medical Center, Kansas City KS, USA
- Alide Tieleman, Radboud university medical center, Nijmegen, the Netherlands
- Monique Plieger, Radboud university medical center, Nijmegen, the Netherlands
- Judith van Engelen-Kanters, Radboud university medical center, Nijmegen, the Netherlands
- Kaneshia Hives, The Ohio State University Wexner Medical Center, Columbus OH, USA
- Andrea Jaworek, The Ohio State University Wexner Medical Center, Columbus OH, USA
- Sarah Heintzman, The Ohio State University Wexner Medical Center, Columbus OH, USA
- Nikoletta Nikolenko, University College London, London, UK
- Charlotte Massey, University College London, London, UK
- Sara Ismail, Stanford University, Stanford CA, USA
- Tina Duong, Stanford University, Stanford CA, USA
- Alyssa Avilez, University of Colorado, Aurora CO, USA
- Talia Strahler, University of Colorado, Aurora CO, USA
- Claire Gilmartin, St George’s University Hospitals NHS, London, UK
- Claire O’Farrell, St George’s University Hospitals NHS, London, UK
- Corinna Wirner-Piotrowski, LMU University Hospital, Munich, Germany
- Maegan Tyrell, BA, University of Iowa, Iowa City IA, USA
- Amy Yotty, PT, DPT, NCS, University of Iowa, Iowa City IA, USA
- Mariah Stechschulte, UC San Diego Health, San Diego CA, USA
- Kristine Negrete DPT, Neurolab 360, Encinitas, CA, USA
- Dennis Fernando, University of California, Los Angeles CA, USA
- Christy Skura, University of California, Los Angeles CA, USA
- Aramide Balogun, Houston Methodist Neurological Institute, Houston TX, USA
- Dora Maldonado, Houston Methodist Neurological Institute, Houston TX, USA
- Hiroto Takada, NHO Aomori Hospital, Aomori, Japan
- Shizuko Taima, NHO Aomori Hospital, Aomori, Japan
- Suzuki Manabu, NHO Aomori Hospital, Aomori, Japan
- Marlon H. Tamayo Muradas, University of Texas Health, San Antonio TX, USA
- Nicholas Dennis, University of Texas Health, San Antonio TX, USA

